# Bloody-Easy: A Novel, Passive, Small-Volume Blood Collection Device for Challenging Phlebotomy Scenarios

**DOI:** 10.64898/2026.02.04.26345604

**Authors:** Luke Sammartino, Maciej Necki, John Kounetas, Andrew Batty, Chris Smith, Finnbar O’Neill, David J. Collins

## Abstract

Despite significant advancements in diagnostic methodologies, the fundamental approach to venous blood collection has remained largely unchanged since its widespread adoption in the mid-20th century. This stagnation poses considerable challenges, particularly in scenarios involving difficult venous access (DVA). The Bloody-Easy (BE) device represents a novel, passive, low-volume blood collection system engineered to optimize phlebotomy outcomes, especially in these challenging clinical contexts. Our prospective, randomized, crossover study involving 90 healthy volunteers demonstrates that BE achieved comparable or superior sample quality while significantly reducing the volume of blood drawn per session. Furthermore, the device garnered substantial positive feedback from both patients and clinicians. BE offers a potentially cost-neutral and low-risk solution for improving blood collection efficiency and patient experience in critical care, emergency medicine, and paediatric settings, where conventional phlebotomy techniques frequently encounter limitations.

## Introduction

Venipuncture serves as a cornerstone of modern medical diagnostics, underpinning an estimated 70% of clinical decision-making processes^1^. However, the instruments and techniques employed for blood sample acquisition have seen remarkably little innovation since the introduction of the Vacutainer system over 70 years ago^2^. This lack of technological evolution in blood collection stands in stark contrast to the rapid and transformative advancements observed in laboratory analytical platforms.

For the majority of the patient population, conventional phlebotomy techniques prove adequate. Nevertheless, an estimated 25% of blood draws are classified as difficult^3^, often characterized by factors such as poor venous access, uncooperative patient demographics (e.g., paediatric or agitated patients), or the presence of fragile vasculature. In these challenging instances, the ramifications of failed or repeated venipuncture attempts extend to delayed diagnoses, increased healthcare expenditures, and significant patient distress. Moreover, in high-acuity environments like intensive care units (ICUs), the necessity for frequent and often large-volume blood draws can precipitate iatrogenic anemia, a phenomenon with demonstrable negative impacts on patient recovery trajectories and healthcare resource utilization^4^.

Moreover, while the significant negative pressures in evacuated container collection tubes can assist in the reduction of sampling time, this rapid blood flow and residual negative pressures can result in blood damage via the rupture of red blood cells, i.e. hemolysis. This can results in analysis errors, misdiagnosis and need for repeated testing for some analytes, with falsely elevated levels of potassium, lactate dehydrogenase, aspartate aminotransferase, magnesim and phosphate. Notably, up to 20% of blood samples may be hemolyzed during routine sampling^5^, highlighting the significance of this longstanding issue.

The Bloody-Easy (BE) device is a novel, passive-flow blood collection system specifically engineered to mitigate the aforementioned limitations inherent in traditional phlebotomy. Designed as a synergistic adjunct to current clinical practice, particularly in complex scenarios, BE aims to provide a solution that prioritizes precision, patient safety, and comfort without necessitating disruptive alterations to established laboratory workflows.

## System principles

The BE system collects blood in conventional sample tubes. Uniquely, blood flow is generated via positive venous pressures, rather than via negative pressures in a sample collection tube. The BE is further designed to be used with conventional blood sampling needles (e.g. butterfly needles) and deposit samples in up to 4 collection tubes sequentially, with attachment geometries permitting the use of conventional 2-10 mL collection tubes (13, 16 mm diameters) and 0.5 mL microtainers. Figure 1 shows the BE device design, assembly and operation, where Fig. 1a displays the assembled BE device with 4 collection tubes inserted. Fig. 1b shows the BE assembly, with a rotating top component, o-rings, BE device bottom with air-channels and sample tube connectors, into which a variety of tube diameters can be inserted. Sequential sampling is produced via the rotational motion of the BE device top component, creating separate continuous fluidic pathways between the blood sampling tubing and each sample tube (Fig. 1c).

**Figure 1.**
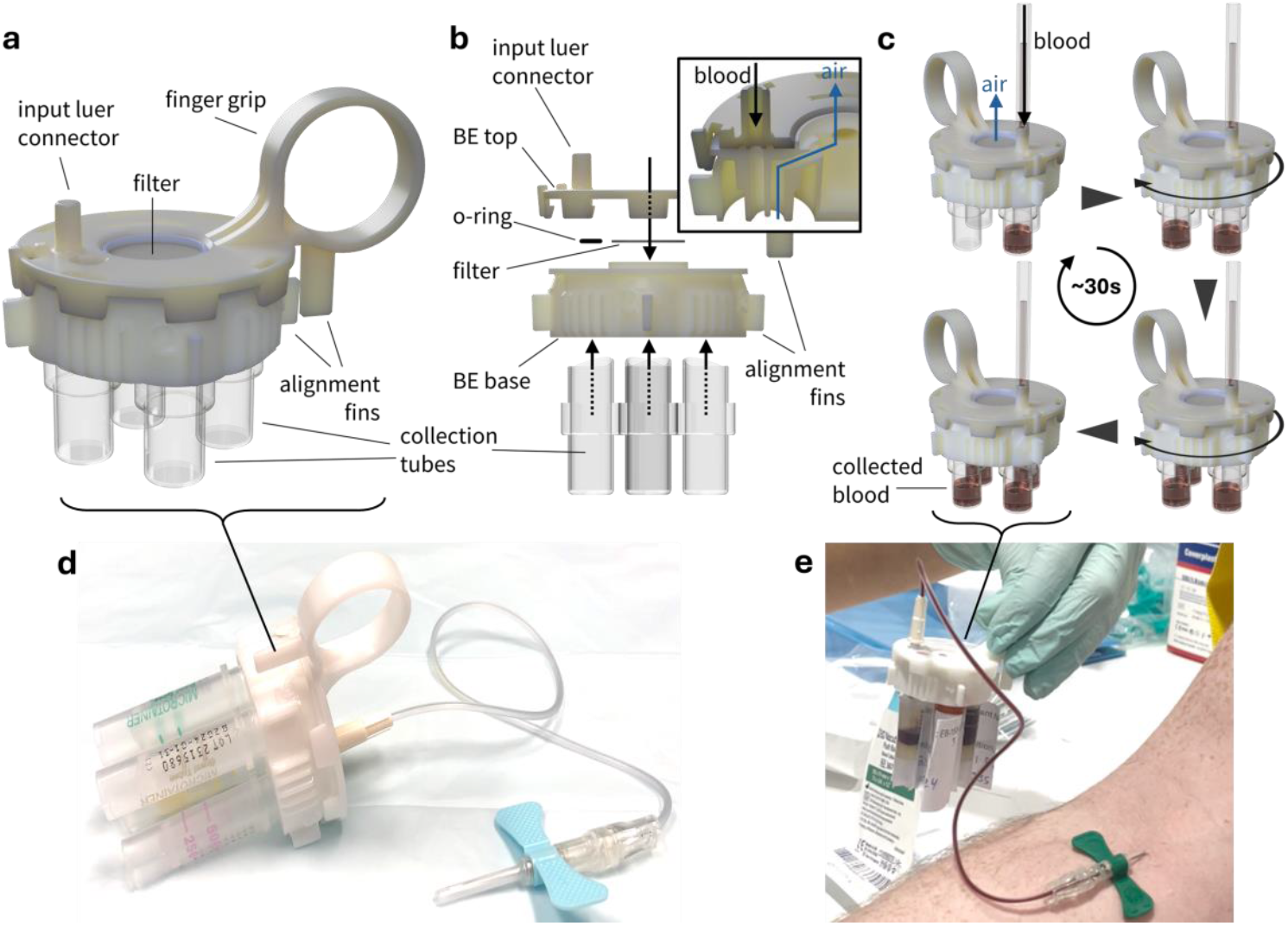
Bloody Easy (BE) device concept. (a) Device. (b) Assembly. (c) Operation. (d) fabricated BE device with butterfly needle collection assembly. (e) Blood collection in microcollection tubes.

While the overall process is broadly similar to conventional blood sampling, with collection of blood sample in collection tubes, there are important differences in the fundamentals and operation of the BE that make this uniquely suited to the sampling of certain patient demographics (e.g. pediatric, elderly).

Importantly, the collection process and filling time is a function of the tubing used, the venous pressure conditions, and height differentials between the sampling location and collection elevation. The viscosity of blood is assumed to be constant at 0.004 mPa·s, with a venous pressure of 10 mmHg. Fig. 2 examines the sensitivity of the flow rate and filling time as a function of these parameters, where the flow rate in the laminar flow regime is given by the Hagen-Poiseuille equation, with

**Figure 2.**
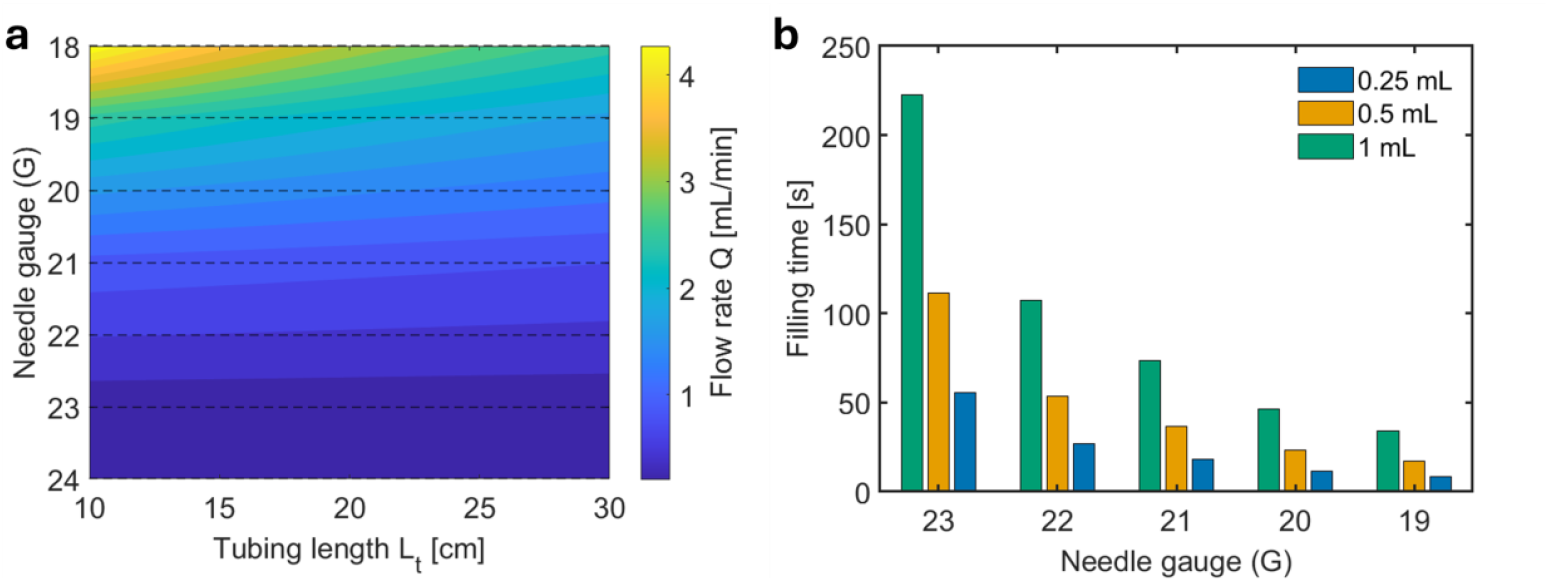
Impact of system parameters on BE filling. (a) Flow rate as a function of tubing length and needle gauge. (b) Filling time as a function of collection volume and needle gauge.

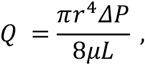

where r is the tubing radius, *ΔP* is the pressure differential, *μ* is the viscosity, and *L* is the length of tubing. Putting this in terms of the tubing diameter *D*, and accounting for the separate length and diameter of the tubing and needle, this yields

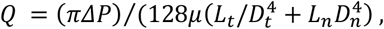

where the subscripts *t* and *n* refer to the tubing and needle, respectively. Notably, Fig. 2 highlights the singular importance of needle gauge to flow rate and filling time, where despite the relatively short length of this needle section, this impacts these parameters to a far greater extent than tubing length due to its significantly smaller internal diameter (e.g. 0.41 mm for a 22G needle vs 1.2mm for typical silicone collection tubing). Accordingly, Fig. 2a shows the dependence of these parameters on flow rate, with an 18G needle resulting in a ∼6x increase in flow rate compared to a 22G needle for a 15 cm tubing length, though the needle component impacts the flow rate proportionally less for increasing tube length. Fig. 2b further demonstrates the impact of needle size on filling time for volumes of 0.5-2mL (30 cm tubing length, 19-23G), where larger needle diameters equate to much shorter filling times. Notably, fill volumes for typical microtainers in clinical testing are between 250-500 µl, making filling times more practical for such lower collect volumes^6^.

## Engineering & Operation

To best suit blood sampling among the particularly difficult patient demographics, BE has been engineered to conform to three core system requirements that we have identified as critical to improve upon the currently gold standard methods. Firstly, both patient and medical staff experience had to be improved. BE features a uniquely rotating hub that is actuated by one handed operation. With one finger securely holding the device by the molded index finger ring, the thumb and middle finger are used to rotate the hub to align the blood channels and enable flow. A considerable number of design iterations has been spent on ensuring that both the size of the ring as well as the position of alignment grooves enables an easy and intuitive user experience. While not recommended, thanks to the alignment features of the rotating hub users can cycle through samples without taking their eyes off the patient. Ultimately, this design characteristic enables medical staff to retain a free hand that can be used for tasks such as securely taping down the collection needle, comforting or holding down the patient, or used for gestures that can make any conversation appear more natural and take away the stress of the collection procedure.

Secondly, a dramatic reduction in blood sample volume alongside an improvement in sample quality had to be achieved. Notably, BE utilizes native venous pressure, between 8-12 mmHg (1.1-1.6 kPa)^7^, to drive blood flow. This compares to a minimum sample container pressure of ∼150 mmHg (20kPa) below sea level atmospheric pressure (101.3 kPa), representing more than an order of magnitude greater pressure differential with the use of an evacuated container. Accordingly, given the scaling between pressure and flow velocity in laminar, Hagen-Poiseuille flow, this results in an increased sampling rate for a given fluidic pathway (needle + tubing), albeit at the expense of producing non-physiological, high-shear rate flow for vacutainer collection. Moreover, this flow rate will decrease over the course of sampling as the blood volume inside the vacuum container increases, as compared to the constant and physiological flow rate and shear driven by venous pressure as in the BE device. The BE device is able to remove displaced air from sample tubes via the use of air-channels, shown in Fig. 1c. These are channels have been designed to meet at a common outlet-reservoir feature. This facilitates space for handling any sample overflow that would collect in the overflow cavity of the device that is further capped by a filter.

The final requirement of BE is to seamlessly integrate with the existing pathology practices and workflows. The rotating hub has been designed to accept most of the commonly used blood collection tubes. Each port on the hub features a set of features that allows tubes of various common diameters to securely fasten within the ports. This way BE can be pre-loaded with a selection of collection containers for rapid deployment during the patient’s visit. To maintain container and sample sterility, an adhesive polymer filter is utilized on the common air channel outlet, preventing the ingress of contaminants, as well as the egress of any excess sampled blood that exceeds the volume of the collection container. To aid the ease of distribution and acquisition, BE devices are designed to be manufactured at a low cost. The two main plastic parts of the device are manufactured by cost effective molding process, while the only two other parts – blood passage O-ring and the polymer filter – are sourced from readily available distributors and require no special tools to fit. The overall device is assembled by snap fit between the rotating hub and the receiver and disposed of after use.

### Results

Fig. 3 shows the collection parameters and key outcomes, highlighting spoilage rates (clotting or haemolysis) in samples obtained in the BE device vs. the vacuum collection system. While the total volume obtained is more than an order of magnitude smaller in the BE device, the collection tubes are distinct; whereas the BE device utilises microcollection tubes, the vacuum system approach typically requires larger volumes for consistent operation. Nevertheless, both volumes are sufficient for clinical analysis^6^. Both blood collection methodologies demonstrated efficiency within clinically acceptable timing thresholds. Importantly, the BE device consistently achieved target fill volumes within ∼30 seconds, an appropriate timeframe for passive flow systems to minimize the risk of pre-analytical clotting,

**Figure 3.**
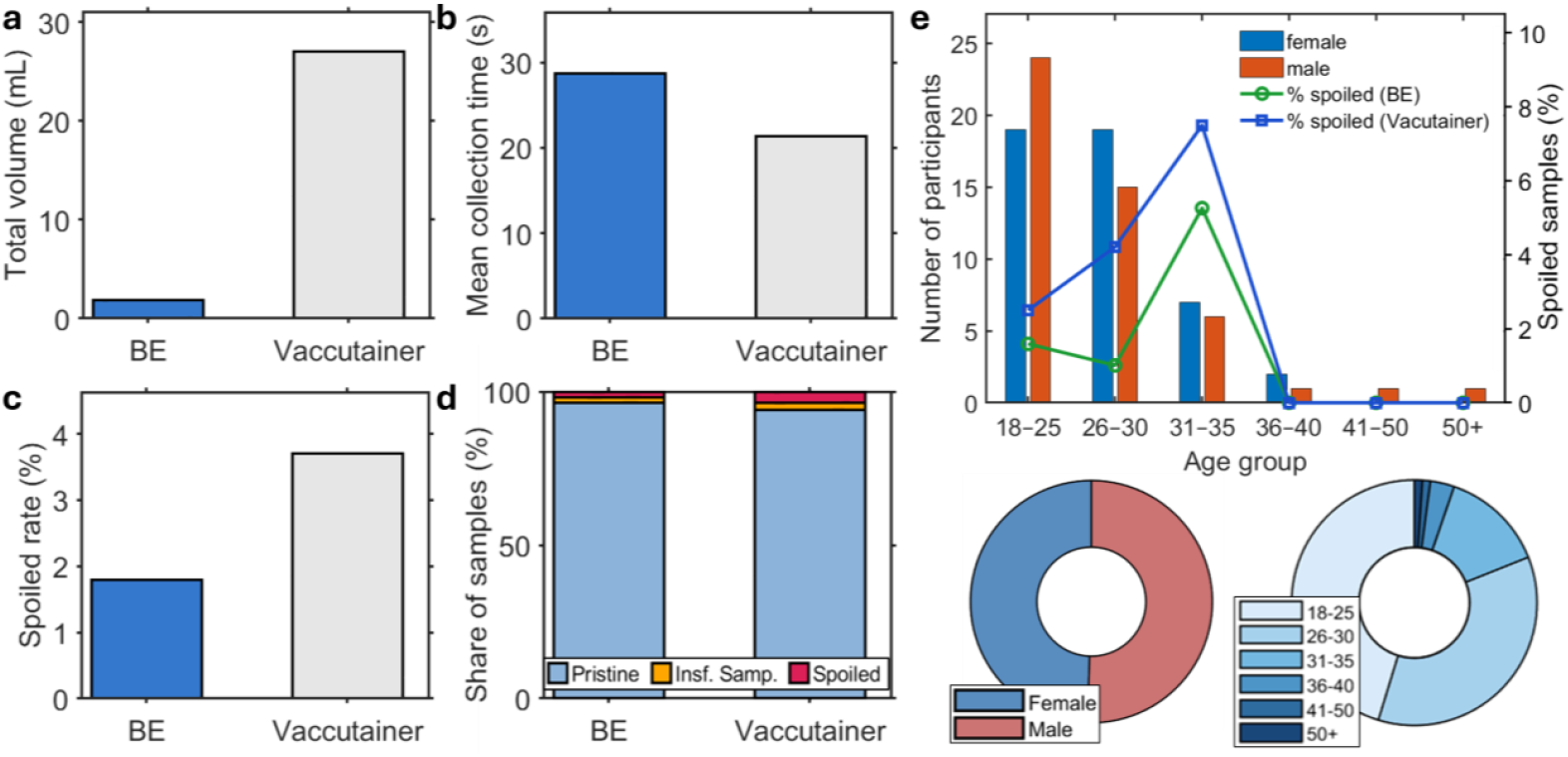
Performance of the BE device compared to vacutainer systems. (a) Average volume collected during phlebotomy over given (b) mean collection times for BE and vacutainer systems (270 BE samples, 278 vacutainer samples, n = 95 participants). (c) Spoilage (clotting or haemolysis) rates for both systems, as well as (d) fraction of each including cases where insufficient samples were collected for analysis. (e) Age cohort distribution (18–25: 45.3%, 26–30: 35.8%, 31–35: 13.7%, ≥36: 5.4%) with near-equal sex representation (47 female, 48 male; n = 95). Plot shows the overlaid spoilage rate by cohort.

**Figure 4.**
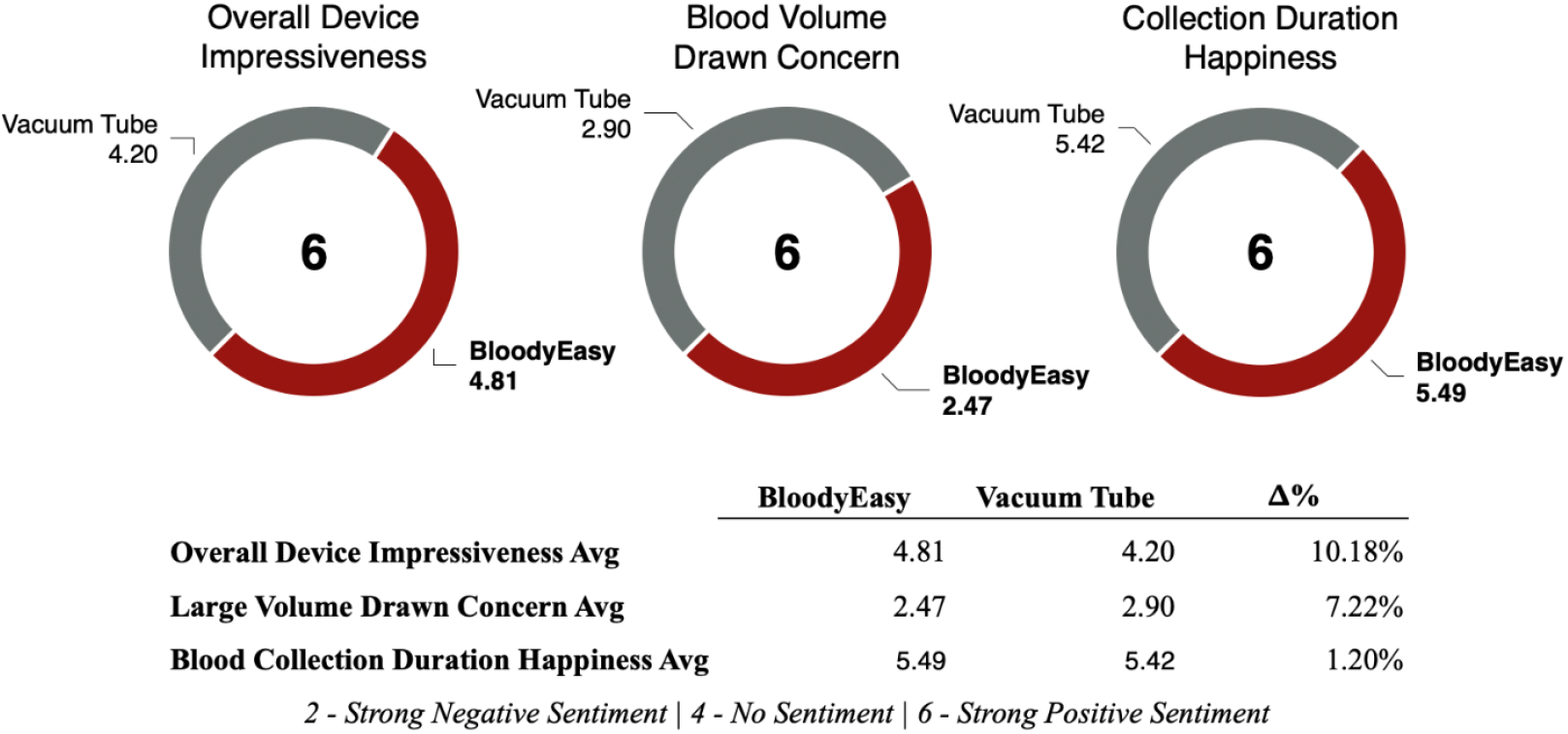
Survey data results, with questions on device impressiveness, concern over blood volume collection, and collection duration.

Analysis of sample quality revealed a markedly lower incidence of quality issues with samples collected using the BE device. Specifically, only 1.8% of BE samples were deemed spoiled, including haemolysed and clotted samples, in contrast to 3.7% of vacuum system samples. Fig. 3d further shows the total fraction of successful collects, including 1.8% and 2.2% of samples where an insufficient volume was collected, indicating insufficient blood flow to obtain a sample. Fig. 3e shows the demographic composition of the participant cohort, with 94.7% being 35 or less. Interestingly, excluding the 5.3% of participants >35, spoilage rates increase for both collection approaches with age, though the spoilage rate with the BE device is universally lower.

### User Feedback and Clinical Utility

#### Patient Feedback

A significant majority of patients (90%) reported that the BE device offered **equal or superior comfort** compared to traditional venipuncture. Elderly and paediatric patient cohorts particularly favored BE, especially when non-traditional or challenging venous access sites were utilized.

In isolation, both devices made a similar impression on the participants in terms of their impressiveness. While the measure is subjective, the participants were able to observe the blood collection process with use of each device. As such they were subject to forming opinions on device handling, perceived ease of use by the clinician, able to observe blood flow into the device, and the technique employed to complete the blood collection in a timely manner. On each of the measures the BE device received marginally improved metrics compared to the vacuum system, including device impressiveness and subjective concern over blood collection volume. For blood collection duration, both devices received almost identical average scores from the participants, demonstrating that the BE device is perceived to perform equally fast as the vacuum tubes.

Fig. 5 shows representative results from a typical metabolic panel, showing equivalent results for sodium, potassium, chloride, bicarbonate, urea, creatinine, and glucose, demonstrating equivalent clinical laboratory testing outcomes between BE and vacuum collection systems. Notably, the measured outputs are substantially impacted by the presence of hemolysis, where these values reflect either a misread if used clinically, or require the need for a subsequent redraw to obtain an accurate measurement. The hemolysis rates for each system are presented in Fig. 5h.

**Figure 5.**
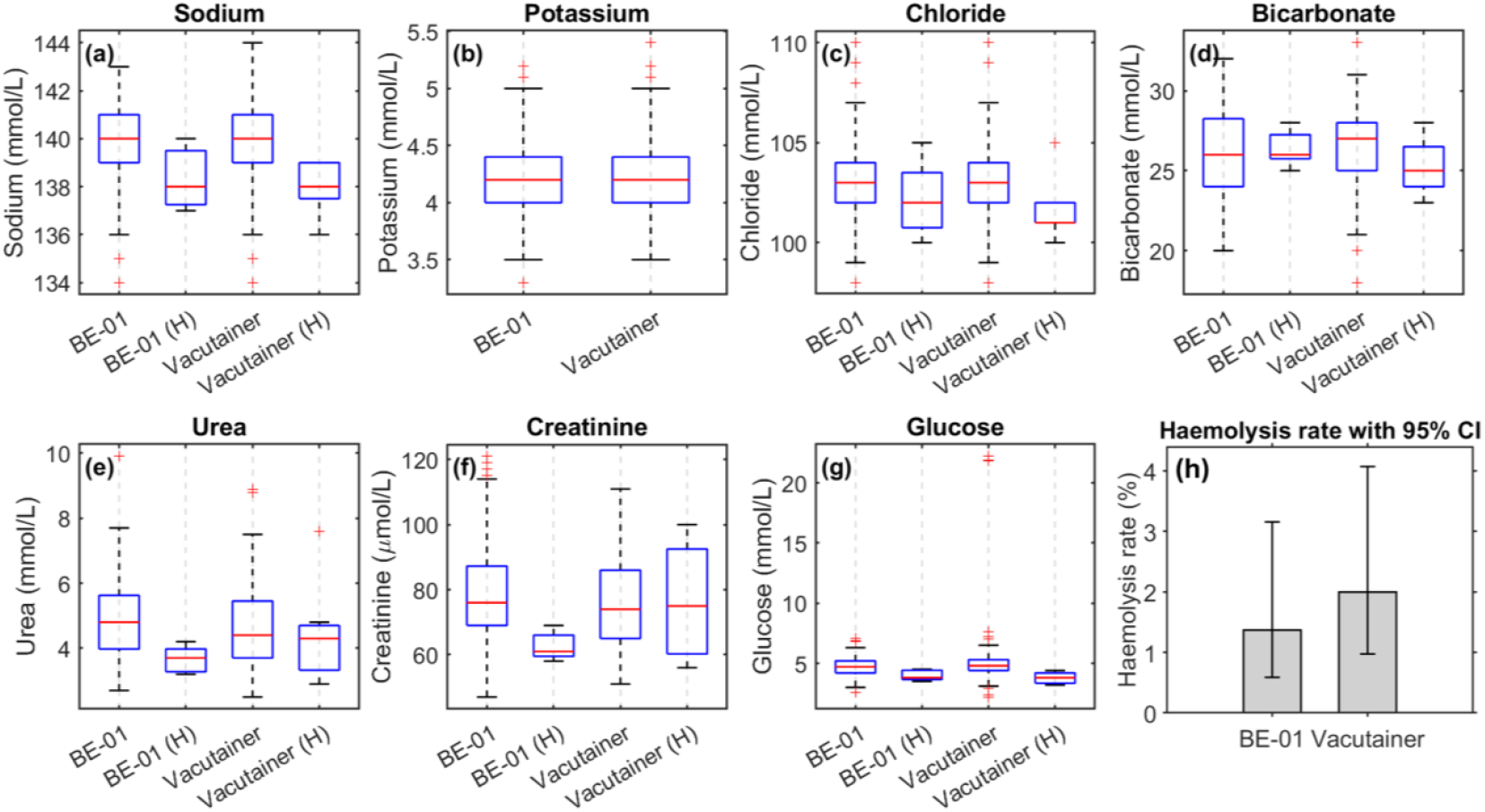
Clinical analysis results. Bloody easy device (BE-01) and vacuum system (vacutainer) analysis results across all n=95 patients for each system. (H) indicates a hemolyzed sample. (a) Sodium, (b) potassium, (c) chloride, (d) bicarbonate, (e) urea, (f) creatinine, and (g) glucose results. (h) hemolysis rate with 95% confidence intervals for BE and vacuum systems.

#### Phlebotomist Feedback

Trained phlebotomists expressed a distinct preference for BE when encountering patients with fragile veins or those who were uncooperative. They specifically highlighted the improved control and reduced incidence of needle dislodgement when using the butterfly-only configuration with BE.

#### Clinician Feedback

Clinicians underscored the significant clinical value of BE’s capability to collect blood from existing indwelling intravenous catheters without compromising line integrity. Renal and oncology specialists, in particular, voiced strong interest in the BE device for chronic care scenarios, citing its potential to minimize repeated venipunctures in their patient populations.

## Discussion

The Bloody-Easy device represents a meaningful and timely innovation in the field of phlebotomy, specifically addressing the persistent limitations of standard vacuum-based techniques in challenging clinical scenarios. We demonstrate that the passive mechanism of BE intrinsically reduces the risks associated with haemolysis, reducing its incidence by an order of magnitude in a controlled study. Vein collapse, particularly in elderly patients, was reduced, as evidenced by the reduction in difficulty in obtaining blood draws. While the venous pressure used to passively drive flow results in a lower flow rate, necessitating the use of smaller sample collection tubes for equivalent collect times as compared with evacuated sample containers, the BE device most suited to patients who are particularly prone to collapse, or for critical pathology tests that are sensitive to the presence of haemolysis. Moreover, the small, ∼0.5 mL sample sizes obtained in BE-compatible microtainers evidence the ability to reduce the risk **of iatrogenic anemia** in patients who may be routinely sampled, particularly crucial in environments necessitating frequent blood draws.

The haemolysis rates for the trial across all devices, including our control method, is substantially lower than reported in the industry. This could be due to several factors, such as the specimens all being collected by a professional phlebotomist (instead of ward assistants), using larger volume specimen tubes. Another potential factor could be due to the average of participants in the trial (being predominantly in their 20-30s) being significantly lower than the median age, which could be due to the increased difficulty of blood collects in older patients resulting from practitioners having difficulty accessing their veins.

A key advantage of BE is its seamless compatibility with existing capillary tube infrastructure, obviating the need for costly or complex downstream laboratory modifications. Furthermore, its intuitive operational interface, coupled with its use of standard butterfly needles, ensures a rapid operator uptake and effortless integration into current clinical practice.

From an economic perspective, BE demonstrates a cost-neutral advantage. Whereas the widespread adoption and availability of evacuated container based sample collection means that this is likely to remain the dominant collection method, the synergistic use of BE in patient demographics that are poorly served by this approach (e.g. elderly, paediatric, ICU patients) is particularly promising. Even with a conservative adoption rate (e.g., 5–10% of total draws), the device’s benefits are projected to offset costs associated with recollection procedures, patient distress, and clinical delays in theses patient groups—factors that impose substantial burdens on healthcare systems. Its demonstrated ability to safely and effectively collect blood from pre-existing IV lines further offers a considerable benefit in time-critical environments such as emergency departments and ICUs, where maintaining vascular access is paramount.

### Limitations

This study was conducted at a single center, which inherently limits generalizability and introduces potential for limited operator variation. Furthermore, the study design necessitated an unblinded approach due to the visible differences between the BE device and the standard vacuum system. Future multi-centre studies involving a broader range of operators and diverse patient populations, particularly within high-acuity and paediatric hospital settings, are warranted to further validate these findings and explore the full clinical utility of the BE device.

## Conclusion

The Bloody-Easy device is a simple, safe, and highly effective tool for optimizing blood collection in challenging scenarios. Its passive, low-volume approach offers dual protection for both the integrity of the collected sample and the well-being of the patient, while simultaneously ensuring seamless integration into current phlebotomy workflows. By providing an enhanced solution for blood collection in the most vulnerable and complex patient cases, BE addresses a long-standing and critical unmet need in contemporary clinical care.

## Methods

### Study Design and Ethical Considerations

A prospective, randomized, crossover study was conducted at the University of Melbourne Department of Biomedical Engineering. Institutional Ethics Committee approval was secured prior to study commencement (Reference: CT28666). A total of 90 healthy volunteers were enrolled, including paediatric subjects for whom appropriate parental informed consent was obtained in accordance with ethical guidelines.

### Phlebotomy Procedure

Each participant underwent two distinct phlebotomy sessions, precisely one week apart to minimize carry-over effects. In each session, a standardized set of four blood collection tubes (two EDTA tubes for haematology assays and two Lithium Heparin tubes for biochemistry analysis) were obtained using either the **BE device** or the **standard vacuum-assisted collection system**. The assignment of collection method for the initial session was randomized, and the techniques were subsequently reversed for the second session (crossover design).

The BE device employed a **passive-flow collection technique** utilizing 21 gauge butterfly needles connected to standard capillary collection tubes. In contrast, control samples were acquired using conventional Vacutainer systems, which rely on a pre-determined vacuum for blood aspiration. The target blood volume for each BE session was approximately 1.9–2.0 mL, significantly lower than the approximate 27 mL typically drawn during a standard vacuum collection session for the equivalent four tubes.

### Outcome Measures

The primary and secondary outcome measures assessed during the study included:

- **Sample collection time:** The total duration required to successfully fill the four designated blood tubes.
- **Number of attempts per draw:** The count of venipuncture attempts required to achieve successful blood flow.
- **Sample quality:** Categorization of collected samples as pristine, mildly tainted (e.g., subtle haemolysis), or spoiled (e.g., gross haemolysis, clotting, insufficient volume).
- **Operator and participant feedback:** Qualitative assessment of device usability, comfort, and preference through structured questionnaires and interviews.
- **Device performance with indwelling IV catheters:** A pre-defined subset of participants assessed the BE device’s efficacy and safety when collecting blood from existing intravenous access lines without compromising line integrity.
- **Transport and laboratory processing:** All collected samples were transported and processed by Austin Pathology adhering to their standard operating procedures, ensuring realistic assessment of sample suitability for routine diagnostic analysis.

Sample integrity was rigorously evaluated both visually by trained laboratory personnel and through automated metrics where applicable, using conventional pathology protocols for full blood examination and basic biochemistry.

### BE device Manufacturing

For the clinical study, we utilized 95 BE devices, with one device per patient, manufactured using a vacuum casting method with a white polyurethane amorphous resin material (ARPTECH, Australia). The manufactured BE devices were packed and sterilized, using an **Ethyl Oxide** sterilization process (Sabre Medical, Australia).

## Data Availability

All data produced in the present study are available upon reasonable request to the authors

## Acknowledgements

The authors extend their sincere gratitude to the University of Melbourne Department of Biomedical Engineering for their invaluable support, Austin Pathology for their expert laboratory services, the Romar and Arptech engineering teams for their technical contributions, and all study volunteers for their participation. Special appreciation is also extended to the phlebotomy staff for their critical feedback and dedicated involvement throughout the study.

